# Financing Vaccines for Global Health Security

**DOI:** 10.1101/2020.03.20.20039966

**Authors:** Jonathan T. Vu, Benjamin K. Kaplan, Shomesh Chaudhuri, Monique K. Mansoura, Andrew W. Lo

## Abstract

Recent outbreaks of infectious pathogens such as Zika, Ebola, and COVID-19 have underscored the need for the dependable availability of vaccines against emerging infectious diseases (EIDs). The cost and risk of R&D programs and uniquely unpredictable demand for EID vaccines have discouraged vaccine developers, and government and nonprofit agencies have been unable to provide timely or sufficient incentives for their development and sustained supply. We analyze the economic returns of a portfolio of EID vaccine assets, and find that under realistic financing assumptions, the expected returns are significantly negative, implying that the private sector is unlikely to address this need without public-sector intervention. We have sized the financing deficit for this portfolio and propose several potential solutions, including price increases, enhanced public-private partnerships, and subscription models through which individuals would pay annual fees to obtain access to a portfolio of vaccines in the event of an outbreak.

## Introduction

In this study, we examine the economic feasibility of developing and supporting a portfolio of vaccines for the world’s most threatening emerging infectious diseases (EIDs) as determined by scientific experts, drawing from the list of targets made by the recently launched global initiative, the Coalition for Epidemic Preparedness Innovations (CEPI) *(1– 3)*. Our portfolio is composed of the 141 preclinical assets identified by Gouglas *et al*. to be targeting the priority diseases.

The risks of EIDs are inherently dynamic and largely unpredictable. New threats persist, including the recent outbreak of a novel coronavirus COVID-19 emerging from Wuhan, China *(4)* Government leaders face formidable decisions about the provision of health security measures against outbreaks of these threats. Global actors are seeking to diminish the danger that these pathogens pose to the wellbeing of nations, regions, and the world. Given the range of potential biological threats, their unpredictability, and the limited resources available to address them, policymakers must necessarily prioritize their readiness efforts based on limited knowledge. All too often, they are forced to choose between priorities, and construct so-called limited lists of treatments, using testimony from teams of experts to inform these decisions. As history has shown, however, this approach leaves society vulnerable to unforeseen outbreaks. Therefore, a more rational approach is to develop a broad portfolio of vaccines in a coordinated manner, mitigating the future risk posed by unpredictable outbreaks of these diseases.

Uncontrolled outbreaks of EIDs, defined as infections that have “recently appeared within a population, or those whose incidence or geographic range is rapidly increasing or threatens to increase in the near future” *(5)*, have the potential to devastate populations globally, both in terms of lives lost and economic value destroyed. Notable recent outbreaks of EIDs include the 1998 Nipah outbreak in Malaysia, the 2003 SARS outbreak in China, and the 2014 Ebola outbreak. In addition to the thousands of lives lost, the economic costs of these outbreaks are estimated as $671 million, $40 billion, and $2.2 billion, respectively *(5–8)*.

As the world becomes more globalized, urbanized, and exposed to the effects of climate change, the danger of infectious diseases has become an even greater concern *(9)*, as emerging and re-emerging strains become more diverse, and outbreaks become more frequent. While distinct from the emerging infectious diseases, influenza serves as the best example of the destruction that viruses with pandemic potential can inflict on the modern world. As a baseline, avian influenza outbreaks in the U.S. since late 2014 have caused economy-wide losses estimated at $3.3 billion domestically, and have significantly disrupted trade *(10)*. The 1918 influenza pandemic, however, is estimated to have infected 500 million people and killed 3-5% of the world’s population. In 2006, Dr. Larry Brilliant stated that 90% of the epidemiologists in his confidence agreed that there would be a large influenza pandemic within two generations, in which 1 billion people would sicken, 165 million would die, and the global economy would lose $1 to $3 trillion *(11)* (see Supplementary Materials for further discussion). Controlling EIDs before they have the chance to reach comparable scale represents a significant opportunity to prevent similar loss.

Despite the threat that these diseases pose to global health and security, however, there are few economic incentives for manufacturers to develop preventative vaccines for EIDs, due to the high costs of R&D and the uncertain future demand. Even if protection against these emerging diseases were immediately achievable with existing technology, development costs are significant *(12)*, as they are for any pharmaceutical development program. Pronker *et al. (13)* estimate that it costs between $200–900 million for a new vaccine to be created. Failure to gain approval also poses a substantial risk, as successful passage through clinical trials only occurs 6–11% of the time *(13, 14)*. Regulatory challenges are particularly prominent in EID vaccine development, as viable candidates are rarely available for distribution during outbreaks, making safety and efficacy testing difficult. As a result, vaccine development for EIDs has been reactive and technologically conservative *(15)*.

In spite of these substantial difficulties—or perhaps because of them—new global initiatives have drawn attention to the need for new approaches to encourage the development of vaccines against EIDs *(16, 17)*. International collaborations like CEPI have drawn extensive public, private, NGO, and academic attention to the perils of global epidemic unpreparedness *(18)*.

This crisis-driven expanded interest in vaccines to address epidemic threats is encouraging, but there is still much work to be done. There needs to be a viable, sustainable business model that will align the financial incentives of stakeholders to encourage the necessary investment in vaccine development *(19, 20)*. While governments and international agencies have striven to create incentives to attract additional private sector investment in vaccine development, these efforts have so far failed in attracting sufficient capital to enhance preparedness against the world’s most deadly emerging pathogens *(21)*.

Several mechanisms have recently been proposed or implemented to create incentives for industry to develop vaccines and other medical countermeasures for EIDs *(22)*. Beyond the “push mechanism” of significant R&D support, these mechanisms provide some measure of a “pull incentive,” recognizing that traditional market forces are insufficient to secure global health security aims. These strategies include the direct government acquisition of stockpiles of vaccines, the use of prizes, priority review vouchers, and the establishment of advance market commitments, each of which is described in more detail in Supplemental Materials. However, to date, none of these strategies have been deemed to be effective in addressing the growing threat of EIDs.

Previous research has demonstrated that a novel ‘megafund’ financing strategy is capable of generating returns that could attract untapped financial resources to fund the development of a portfolio of drug development programs *(23, 24)*. In this study, we address this possibility by simulating the financial performance of a hypothetical megafund portfolio of 141 preclinical EID vaccine development programs across 9 different EIDs for which there is currently no approved prophylactic vaccine. Under current business conditions, we determine a private sector solution for the comprehensive development of EID vaccines is not yet feasible, and quantify the gap so as to inform current policy discussions regarding the need for public-sector intervention.

We conclude with a discussion of three possible solutions to this challenge: 1) establishing a global acquisition fund for EID vaccines, in which governments around the world collaborate; 2) raising the price of portfolio vaccines by two orders of magnitude; and 3) creating a subscription model for vaccines, through which the global at-risk population pays an annual fee to fund the development of and ensure access to a predefined list of vaccines for EIDs.

## Megafund Rationale

To create further incentives for investing in this space, we hypothesize the creation of an EID megafund based on the model developed by Fernandez *et al. (23)*, which uses portfolio theory and securitization to reduce investment risk in these assets. In financial engineering, the practice of securitization requires the creation of a legal entity that issues debt and equity to investors, using the capital raised to acquire a portfolio of underlying assets—in this case, vaccine candidates targeting EIDs. These assets subsequently serve as collateral, and their future cash flows service the debt incurred to acquire them, paying the interest and principal of the issued bonds. Once the debt has been repaid, equity holders receive the residual value. If the portfolio’s cash flows are insufficient to meet the obligations to the bondholders, the collateral will be transferred to bondholders through standard bankruptcy proceedings.

Given the characteristically high risk of default of candidates in the early stages of development, and the need for increased financial investment in vaccine research as a whole, securitization in the form of a vaccine megafund offers several key benefits. The securitization of vaccine research enables investors to reduce their risk of financial loss to a scale that is not readily achievable under current financing mechanisms, as they can invest in many vaccine projects at once, thus increasing the likelihood of at least one success. The normalization of returns created by the construction of an asset portfolio permits the issuance of debt, which allows fixed-income investors to gain exposure in a space that is traditionally too risky to represent a compelling opportunity for investment. The ability to issue debt is critical, because bond markets have much greater access to capital than does venture capital or the private and public equity markets. This allows the megafund to raise enough funding to purchase an array of assets and reach its critical threshold of diversification.

One notable benefit of our megafund approach is that it hedges against the societal risk that the world will not have the ‘right’ vaccine it needs for the next EID outbreak. To date, the U.S. government and CEPI programs have been forced to severely limit their portfolios, due to funding constraints. This approach allows us to assess the opportunity of addressing 9 of the world’s most threatening EIDs at once.

While the megafund approach is effective at reducing the development risk of EID vaccines, it should be emphasized that the success of this technique hinges upon securitizing assets that have the potential to be profitable individually if the development effort is successful. This flies in the face of conventional pharma wisdom that vaccines are commercially challenging, not only because of development risk but also because of the unpredictability of outbreaks and constraints on pricing when outbreaks occur. However, to quantify the gap between reality and commercial viability—and in light of global stakeholders’ ongoing efforts to raise funding to combat these diseases—we suspended belief in this presumption so as to allow the financial analysis to determine the profitability of the EID portfolio in an unbiased fashion. Based on available pipeline data, an analysis by Gouglas *et al*. projects that the cost of progressing at least one vaccine candidate through the end of phase 2a against a comparable portfolio of 11 emerging infectious diseases would cost between $2.8 and $3.7 billion *(3)*. Our approach builds upon this analysis by quantifying the gap between the estimated costs of development and the sort of returns that would need to be generated by such expenditure in order to justify investment.

## Methods

To apply this portfolio approach to EID vaccine development, we began by analyzing the hypothetical investment returns of a portfolio of 141 preclinical EID vaccine development programs across 9 different emerging infections for which there is currently no approved prophylactic vaccine. Our analysis relies on several assumptions and parameters, including estimates of the cost of vaccine development, the length of time from preclinical testing to the filing of a new vaccine license application, the probability of success of each project, and pairwise correlations of success among the projects in the portfolio. The target diseases were selected from CEPI’s Priority Pathogen list, which was based in part upon the WHO’s R&D Blueprint focusing on epidemic prevention *(1, 2)*. We drew our portfolio assets from CEPI pipeline research for each disease on its priority pathogen list *(1, 3)*. (See Supplementary Materials for more details).

The model design is less complicated than that of Fernandez *et al. (23)*. Unlike oncology—a domain with many approved drugs and even more under development—there are currently few EID vaccines available on the market, indicating a paucity of data with which to calibrate our simulations. In setting our simulation parameters, we relied on generic information about the vaccine development process, specific estimates posited by CEPI *(1)*, and qualitative input from scientists with domain-specific expertise.

The present value of out-of-pocket development costs for each of the projects in the portfolio was set to $250 million, based on assumptions made by CEPI about the cost to develop a preclinical asset through phase 2 *(1)*. CEPI further estimates that it will take five years for this development to occur (Figure 1). CEPI proposes that assets at this level of development will justify stockpiling, further development, and conditional usage under emergency conditions, a plan that some experts believe may be feasible *(1, 25)*.

**Figure 1.**
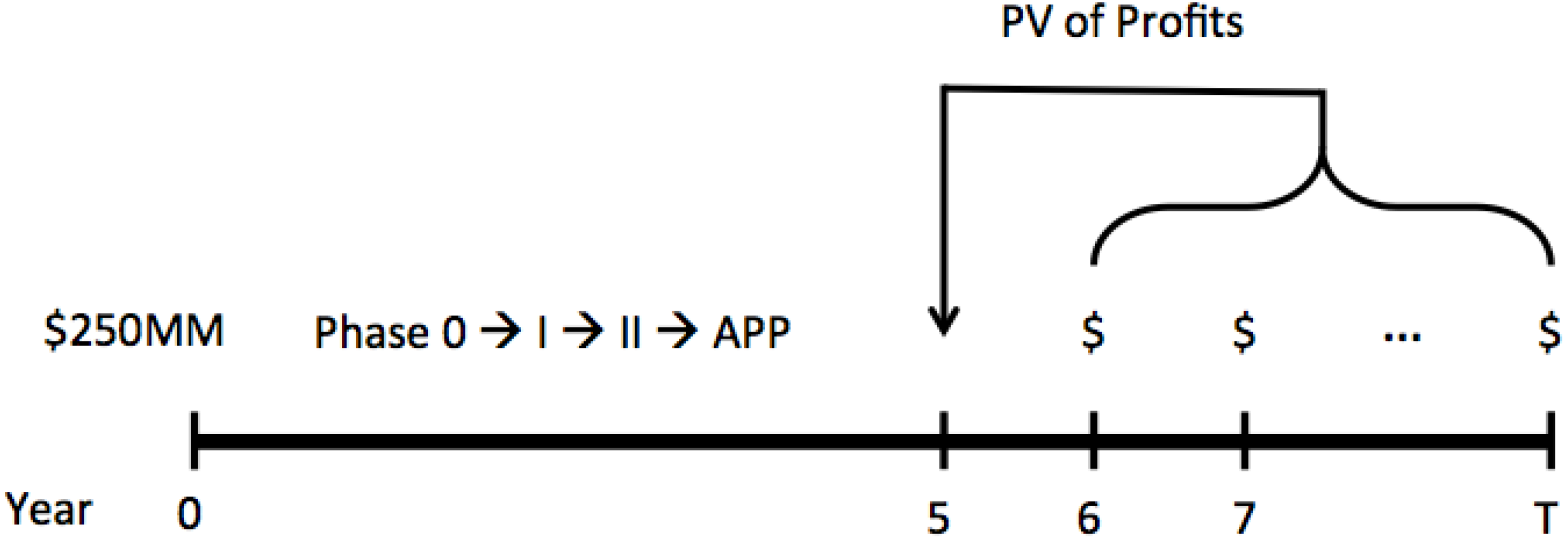
Timeline of a hypothetical EID vaccine development program.

At $250 million per project, a megafund of 141 projects requires $35.25 billion. To determine the returns generated by such a portfolio, we assumed a 15-year period of exclusivity and a 10% cost of capital to calculate the NPV of future cash flows upon approval in year 5. This value must be weighed against the possibility of total loss if the vaccine project fails. An assessment of the megafund’s returns therefore requires estimates of the probabilities of success of each of the 141 vaccine candidate projects as well as the pairwise correlation of success of all possible pairs of assets. The probabilities of success are based on estimates of the compounded probabilities of advancement from preclinical testing to vaccine approval. The probability of development through phase 2 of a vaccine at the start of preclinical testing is 32%, based on the transition probabilities provided by CEPI *(1)*. See Supplementary Materials for details on these estimates as well as the method for assigning pairwise correlations.

Given the inherent unpredictability of a future EID outbreak, we necessarily made several practical assumptions to project revenue. In this model, we assumed that the prophylactic regimen would consist of a single dose of vaccine. The probability of disease outbreak was estimated based on historical outbreaks per disease, while regimen demand was projected using historical outbreak size, potential for pandemic spread, and an assessment of relative clinical severity. These demand parameters were determined respectively by case estimates from documented outbreaks, referencing the Woolhouse assessment for pandemic potential, and comparing the clinical presentation and prognosis for each disease *(26, 27)*. A perceived demand multiplier was assigned based on Woolhouse classification and clinical severity on a five-step scale ranging from mild to severe. The average number of cases and the perceived demand multiplier were used to calculate the number of regimens sold in an outbreak year for each disease. This product, the expected chance of outbreak in a given year based on historic outbreak data, and the expected selling price per vaccine regimen were used to subsequently calculate the annual expected revenue for each disease. The price per regimen was estimated based on whether the disease in question typically affected high-, medium- or low-income countries. The expected price per regimen for each income level was informed by CDC, GAVI, and PAHO vaccine pricing data, respectively *(28–30)*. Please see Supplementary Materials for additional details.

## Results

Table 1 provides estimates of the annual expected revenues from direct sales of vaccines to susceptible populations for the 9 different EIDs considered in the megafund. (Please see Supplementary Materials for more details on how projected revenues were determined.)

**Table 1.**
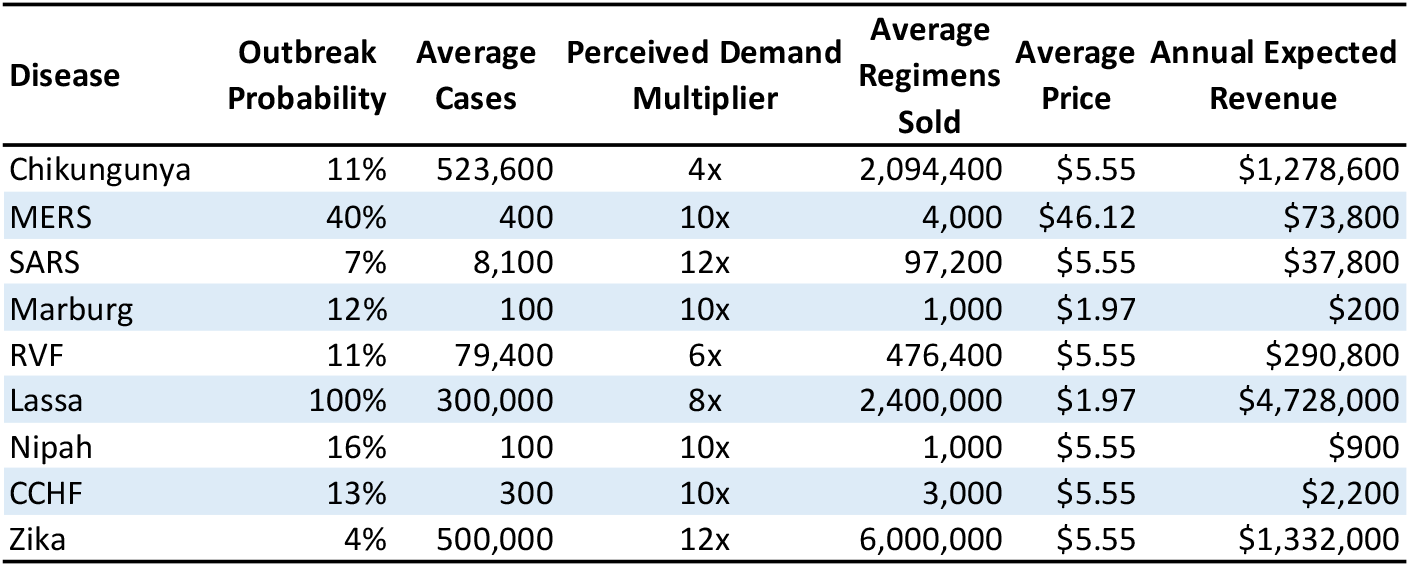
EID vaccine sales. Annual expected revenues from direct sales of vaccines to susceptible populations for 9 different EIDs. All values are annualized.

| Disease | Outbreak Probability | Average Cases | Perceived Demand Multiplier | Average Regimens Sold | Average Price | Annual Expected Revenue |
| --- | --- | --- | --- | --- | --- | --- |
| Chikungunya | 11% | 523,600 | 4x | 2,094,400 | \$5.55 | \$1,278,600 |
| MERS | 40% | 400 | 10x | 4,000 | \$46.12 | \$73,800 |
| SARS | 7% | 8,100 | 12x | 97,200 | \$5.55 | \$37,800 |
| Marburg | 12% | 100 | 10x | 1,000 | \$1.97 | \$200 |
| RVF | 11% | 79,400 | 6x | 476,400 | \$5.55 | \$290,800 |
| Lassa | 100% | 300,000 | 8x | 2,400,000 | \$1.97 | \$4,728,000 |
| Nipah | 16% | 100 | 10x | 1,000 | \$5.55 | \$900 |
| CCHF | 13% | 300 | 10x | 3,000 | \$5.55 | \$2,200 |
| Zika | 4% | 500,000 | 12x | 6,000,000 | \$5.55 | \$1,332,000 |

The simulated investment performance of an EID vaccine portfolio as a function of the commercial potential of each individual vaccine project is provided in Table 2 and illustrated in Figure 2 (please see Supplementary Materials for more information on how returns were calculated). The commercialization potential of these vaccines is consistently very poor, orders of magnitude lower than what would be required to make them commercially viable. The parameter values that are closest to industry averages correspond to the highlighted row in Table 2, in which the expected annual profits upon FDA approval are $1 million, resulting in an NPV per successful EID vaccine of $7.6 million. For these values, the vaccine portfolio’s expected return is −61.1%, with a standard deviation of 4.0%.

**Table 2.** EID megafund risks and returns to investors. Investment returns (%) of a portfolio of 141 preclinical EID vaccine candidates when projects are not independent (with correlation), and when projects are statistically independent (no correlation). The Sharpe ratio is estimated as the ratio of the expected return to the standard deviation. PV(Profits), present value of profits per successful vaccine in year 5; E[R_5y_], expected 5-year return on investment; E[R_1y_], expected annualized return; SD[R_1y_], annualized return standard deviation; CI, confidence interval; SR, Sharpe ratio.

| PV(Profits) |  | With Correlation |  |  |  |  | No Correlation |  |  |  |
| --- | --- | --- | --- | --- | --- | --- | --- | --- | --- | --- |
| in \$MM | E[R<sub>5y</sub>] | E[R<sub>1y</sub>] | SD[R<sub>1y</sub>] | 95% CI | SR | E[R<sub>1y</sub>] | SD[R<sub>1y</sub>] | 95% CI | SR | |
| 0.10 | -100.0 | -83.6 | 1.7 | (-87.4, -80.9) | — | -83.3 | 0.4 | (-84.2, -82.6) | — |  |
| 1.0 | -99.9 | -74.1 | 2.7 | (-80.1, -69.7) | — | -73.6 | 0.7 | (-74.9, -72.4) | — |  |
| 7.60 | -99.0 | -61.1 | 4 | (-70.2, -54.6) | — | -60.4 | 1 | (-62.4, -58.5) | — |  |
| 10 | -98.7 | -58.9 | 4.3 | (-68.5, -51.9) | — | -58.1 | 1 | (-60.3, -56.2) | — |  |
| 100 | -87.0 | -34.8 | 6.8 | (-50.0, -23.9) | — | -33.6 | 1.6 | (-37.0, -30.5) | — |  |
| 772 | 0.0 | -1.9 | 10.2 | (-24.8, 14.5) | — | 0 | 2.5 | (-5.2, 4.5) | — |  |
| 1,000 | 29.7 | 3.3 | 10.7 | (-20.8, 20.6) | 0.3 | 5.2 | 2.6 | (-0.1, 10.0) | 2.01 |  |
| 10,000 | 1197.1 | 63.8 | 17 | (25.6, 91.2) | 3.8 | 66.8 | 4.1 | (58.3, 74.5) | 16.3 |  |

**Figure 2.**
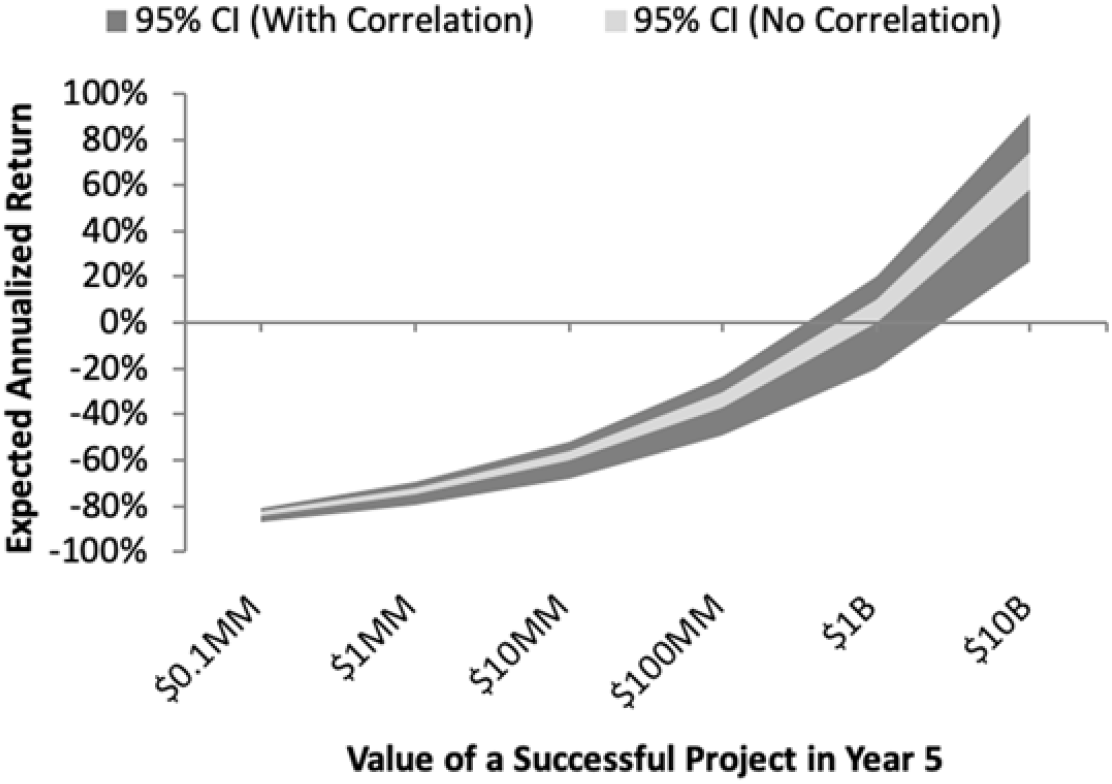
EID megafund risks and returns to investors. Investment returns and risks of a portfolio of 141 preclinical EID vaccine candidates when projects are not independent (with correlation), and when projects are statistically independent (no correlation). Expected returns break even when the annual expected profit per successful project is $772 million. CI, confidence interval.

For completeness, Table 2 also reports megafund performance statistics for several other sets of parameters. The breakeven point, where the megafund’s expected 5-year return is 0%, occurs as the NPV of a successful vaccine reaches $772 million, two orders of magnitude greater than our current estimates using past averages for costs, revenues, probabilities of success and outbreak, and other information. However, for an NPV of $1 billion, the vaccine portfolio becomes marginally profitable, and at $10 billion, it is highly profitable. These results suggest that many of the model parameters would have to change drastically for the portfolio to be profitable. In fact, holding all else equal, simply breaking even would require selling vaccines at approximately 100 times the price assumed in our simulations.

Megafunds are, of course, not the only business model through which vaccines can be developed. Traditionally, large pharmaceutical companies have incorporated vaccine programs into broader and highly diversified portfolios of therapeutics across many indications. To explore this possibility, we estimated the impact on risk and reward of incorporating the EID vaccines portfolio into a hypothetical pre-existing and profitable pharma company. Table 3 contains the estimated expected returns and volatilities of a representative top-10, mid-tier, and small-capitalization pharmaceutical company with and without the base case version of the EID vaccine portfolio. The best-case scenario—in which big pharma adds this portfolio to its existing products—turns an otherwise profitable business into an unprofitable one, losing 8.6% per year on average in shareholder value. The results for mid- and small-cap pharma companies are even worse.

**Table 3.** Simulated performance of a hypothetical representative top-10, mid-tier and small-cap pharmaceutical company with and without the EID vaccine portfolio. Pharmaceutical companies are classified according to their North American Industry Classification System (NAICS) code and their market capitalization each year from 2005 to 2016. Return statistics are averaged within each sub-group to form the expected return and standard deviation estimates. The performance of these representative companies combined with the EID vaccine portfolio is estimated by assuming no correlation with vaccine revenues. Market Cap, average market capitalization in billions of dollars; E[R1y], expected annualized return; SD[R1y], annualized return standard deviation; SR, Sharpe ratio.

| Company Type | Without EID Vaccine Portfolio |  |  |  | With EID Vaccine Portfolio |  |
| --- | --- | --- | --- | --- | --- | --- |
| | Market Cap (\$B) | E[R<sub>1y</sub>] | SD[R<sub>1y</sub>] | SR | E[R<sub>1y</sub>] | SD[R<sub>1y</sub>] |
| Top-10 Pharma | 94.1 | 11.10% | 23.80% | 0.47 | –8.6% | 17.40% |
| Mid-Tier Pharma | 12.9 | 14.30% | 32.90% | 0.43 | –40.9% | 9.30% |
| Small-Cap Pharma | 1.6 | 19.60% | 53.20% | 0.37 | –57.6% | 4.50% |

These results are consistent with the biopharma industry’s trend towards fewer companies willing to engage in vaccine R&D, underscoring the infeasibility of a private-sector EID vaccine portfolio given current cost and revenue estimates, and the need for some form of public-sector intervention. A sensitivity analysis of these results to perturbations in our model’s key parameters is provided in the Supplementary Materials. We find that the EID vaccine megafund remains financially unattractive even under relatively optimistic cost and revenue assumptions, implying the necessity for some form of public-sector intervention. These findings may explain the dearth of EID vaccines developed over the past decade.

One intervention is the use of government-backed guarantees to mitigate the downside risk of the EID portfolio. In a guarantee structure, a government agency promises to absorb the initial losses on the portfolio to a predetermined amount, shielding private-sector investors from substantial negative returns. For example, a guarantee on 50% of the portfolio’s principal improves the expected annualized return in the base case scenario from −61.1% to −12.6% (see Table S11 in the Supplementary Materials). While this negative-expected-return scenario is still unlikely to attract investors, expected returns can be further increased using mechanisms such as advance market commitments and priority review vouchers. The guarantee structure—in combination with other existing revenue-boosting mechanisms— has the potential to transform a financially unattractive portfolio of EID vaccine candidates into one that could realistically attract private-sector capital.

Finally, we consider a subscription model under which the largest governments around the world would purchase subscriptions to EID vaccines on behalf of their constituents. To fund the cost of pursuing 141 vaccine targets at $250 million per target (for a total of $35.25 billion), suppose that the governments of the G7 countries agreed to pay a fixed subscription fee per capita over a fixed amortization period to cover this cost. How much would this subscription fee be? For an amortization period of 5 years, and an estimated total G7 population of 770,063,285 (as of 2016, according to the World Bank *(31)*), and a cost of capital of 10%, the per capita annual payment to cover the total cost of $35.25 billion is $12.08 per person per year. If we extend the amortization period to 10 years, the subscription fee declines to $7.45 per person per year. Table 4 contains the per capital subscription fees as a percentage of the annual per capita healthcare expenditure of each G7 country and as expected, the cost is trivial for all countries, ranging from a high of 0.59% for Italy to a low of 0.15% for the US using a 5-year amortization period.

**Table 4.**
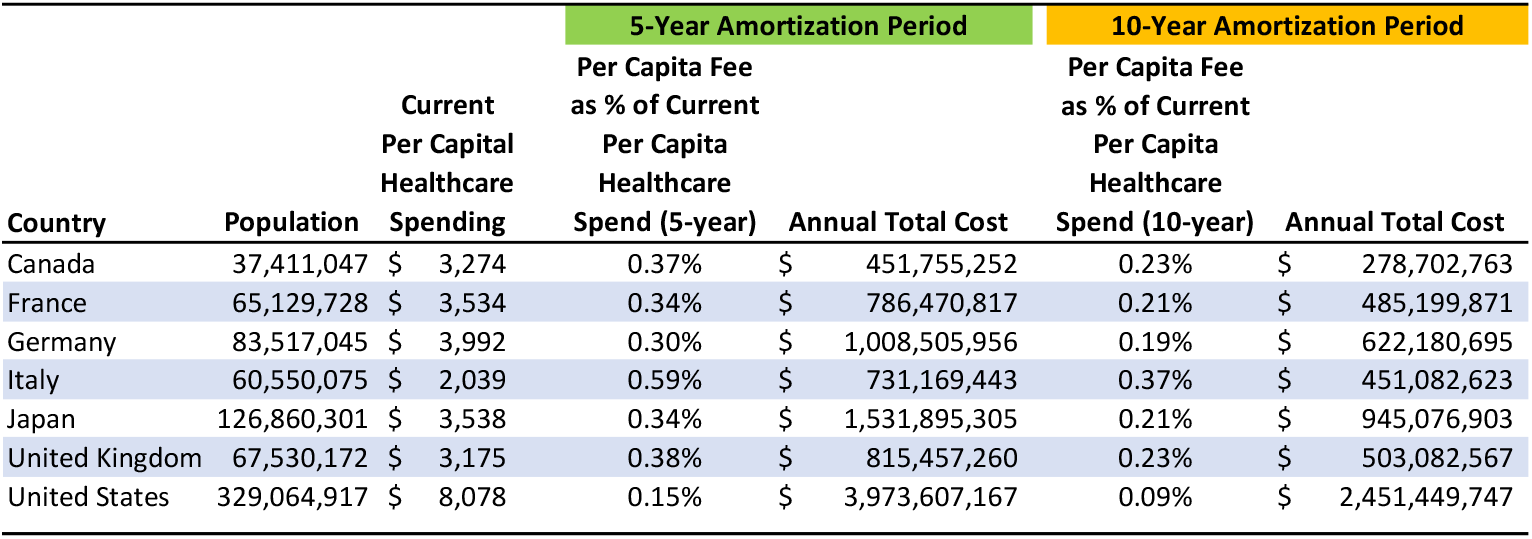
Annual total cost and per capita cost of subscription model for funding a $35.25 billion vaccines development fund by G7 countries where the per capital subscription fee is $12.08 per person per year over a 5-year period or $7.45 per person per year over a 10-year period. Source: authors’ computations based on population and healthcare expenditure data from the World Bank (*31*).

Of course, this subscription model considers only the development cost of vaccines. Once developed, the production and stockpiling of these vaccines would require further funding, but the subscription model can be applied on an ongoing basis, and at a much lower annual cost. Access to these vaccines by non-G7 countries must also be considered, but such access involves political and ethical issues that are beyond the scope of this economic analysis.

These results suggest that a government-led subscription model is financially feasible and would likely yield significant economic and political benefits to all participating governments. While the usual challenges of broad multi-national cooperation must be overcome, early traction from organizations such as Civica Rx suggests that focused, inclusive collaboration can ensure sustained supplies of life-saving drugs *(32)*.

## Discussion

Financing global health security against biological threats remains a persistent challenge. Unfortunately, but not unexpectedly, a weak and uncertain pre-crisis market demand has led to a relative lack of interest in developing vaccines against EIDs. This has left the global community increasingly vulnerable to repeated outbreaks of these viruses. The challenges of EID vaccine development, however, are troubling issues for vaccines more generally. The situation has been described as a crisis, and perhaps rightly so, as there are only four remaining major manufacturers that focus on vaccine development *(25)*.

Vaccines only sell for a fraction of their economic value, in some cases for only a few dollars. They provide myriad benefits, like enabling would-be patients to live longer, healthier lives *(33, 34)*, and bearing yet-undervalued gains in productivity and positive externalities to society at large *(35–37)*. Although the low price of vaccines is meant to benefit individuals and regions with lower incomes, in the long run, it has had the opposite effect, causing them to be medically underserved due to a lack of vaccine investment. Pharmaceutical companies and investors are directing their resources to projects in which the estimated return on investment is more predictable and lucrative. Vaccine prices are currently set far below the prices of drugs that treat other serious conditions, such as cancer, despite the enormous societal value of vaccines in general, and those to ensure global health security in particular. The typical expected risk-adjusted net present value (NPV) of a vaccine in our hypothetical portfolio upon regulatory approval is on the order of only $7.6 million. This is two or three orders of magnitude lower than the comparable value of an approved cancer drug, yet the out-of-pocket costs to develop an EID vaccine are not dissimilar.

In addition to pricing, another challenge lies in assessing the future demand for EID vaccines. Due to the inherent unpredictability in the scale and timing of outbreaks, the future demand for a specific EID vaccine is typically unclear. An additional factor is geopolitical. Diseases that are traditionally found in only a few, lower-income countries may not attract as many R&D dollars because generating a return on investment is more difficult in those limited markets *(25, 38)*. While wealthier governments might issue purchase agreements to assure vaccine sponsors of returns *(38)*, these commitments are more difficult to secure for EIDs in lower-income countries or those undergoing economic hardship. However, an increasing number of stakeholders are realizing the danger of this dynamic for low and high-income countries alike, as under epidemic outbreak conditions, diseases like Zika and Ebola have the potential to spread much further than their traditional locales. The Ebola outbreaks in West Africa in 2014 demonstrate how the absence of vaccine demand prior to an event may result in a tragic loss of life and a regional economic setback. It is a significant concern that years after those outbreaks, the demand for Ebola vaccines remains limited and uncertain, allowing gaps in preparedness to persist *(39–41)*.

Unless these market challenges are addressed, the global population will remain vulnerable to substantial human and economic losses when epidemics and pandemics arise.

We believe that this represents a significant missed opportunity. Aside from the nuclear threat and climate change, pandemics represent one of the most significant existential dangers facing humanity today *(42)*. Nevertheless, investments in preparedness for biological threats remain underfunded, leaving the world vulnerable to catastrophic infectious disease events. With this in mind, we propose several measures that might move the mission for EID vaccine readiness toward financial viability.

Our analysis strongly suggests that reliance solely on private sector investment in EID vaccines is insufficient, given the negative returns achieved by an EID-focused megafund, and the negative impact such a pool of assets would have on an otherwise profitable pharmaceutical company. As a result, if EID vaccine candidates are to be developed, continued private-public cooperation will be imperative, and novel approaches to engage and attract capital will be needed. While bond markets are capable of providing access to substantial amounts of capital to help vaccine development efforts, the resources available to the public sector have great potential as well *(43)*. In 2015, the U.S. spent $9,990 per person on healthcare *(44)*. If we assume that there are 300 million Americans, just 1.25% of this amount of spending would yield $37.46 billion dollars, greater than the projected $35.25 billion it would take to fund the entire EID portfolio of vaccines. While achieving such an allocation of funding would hardly be as simple as this calculation suggests, this thought experiment illustrates that encouraging the development of vaccines that protect against EIDs of pandemic potential is well within the means of the global public and private sector stakeholders, if there is public support and political will. In fact, there is evidence to indicate that people expect and would support further protection from these threats *(45)*.

The U.S. government’s MCM program has demonstrated a capability to create incentives for the development of vaccines that would otherwise not be developed, once sufficient market demand is guaranteed ahead of time. This has been true for anthrax and smallpox as well as for various strains of pre-pandemic influenza, for which the government provides market commitments on the order of $100-200 million per year for successful vaccine development programs *(46, 47)*. While challenges exist (e.g., sustained funding commitments), new initiatives such as CEPI can learn important lessons from these examples *(48, 49)*.

Perhaps key to the problem of EID vaccine funding is a deficiency in the pricing of the risk of infection by EIDs. Although the prevention of epidemics and pandemics saves countless lives and billions of dollars of economic value, the revenue realized by vaccine manufacturers is only a very small fraction of this value. With this in mind, an examination of a capitated fee structure—a subscription model—applied to vaccine development and acquisition is promising. Under the current model, vaccines are purchased a la carte after outbreaks begin.

However, if stakeholders were to pay in advance to develop and stockpile vaccines, viewing their payment as a form of insurance that would maintain epidemic response capabilities and provide protection from EID outbreaks, much like a society-wide immune system, the amount of capital needed to fund these programs might be easier to raise and keep the price per regimen lower. Vaccine developers under this model would most likely sell subscriptions to governments, building upon existing infrastructure, such as the U.S. government’s biodefense and pandemic preparedness programs. To balance the concern that non-subscribers may require vaccine regimens with the objective of encouraging subscription ahead of outbreaks, a tiered pricing scheme rewarding early adoption could be implemented. A private subscription model should also be explored, however, as it would enable individuals, communities, and corporations to take greater ownership in preparedness. Determining precisely who should pay the insurance premium, and who is willing to pay, is essential to this arrangement.

Although this model is a departure from the status quo, promising innovation in vaccine financing is becoming more commonplace. The recent World Bank issue of pandemic bonds and swaps for a Pandemic Emergency Financing Facility (PEF) suggests that when structured appropriately, assets geared toward preparedness can be attractive to investors *(50)*. We believe that our model may shed some light on what will encourage more comprehensive pandemic preparedness by addressing shortcomings in the EID vaccine pipeline.

As demonstrated in our simulations, the investment required to reduce the global risk from EIDs is within reach. Securing these resources, however, will require governments to strengthen their commitments to supporting EID vaccine markets, in order to allow private sector stakeholders and untapped capital to engage with these markets substantively. The recent developments around Sanofi Pasteur’s Zika collaboration highlight the risks of a variable commitment to preparedness. Due to changing epidemiology and internal disputes over potential product pricing, BARDA and Sanofi have chosen to halt further development of their Zika asset, leaving society vulnerable to future outbreaks *(51)*.

As cases like this suggest, government buy-in is integral for long-term pipeline sustainability. Governments can catalyze outside investments through a range of strategies, including guaranteed commitments. Fifteen years of guaranteed revenue via purchase commitments, similar to the U.S. government’s purchase of smallpox and anthrax vaccines, would do well to encourage development efforts. For example, an annual purchase commitment of $150 million per successful vaccine candidate would represent an NPV of $1.14 billion, exceeding our modeled breakeven NPV of $772 million. Our results suggest that investment in this space is highly unattractive to the private sector, requiring commitments of the aforementioned magnitude for development viability; as highlighted above, either the price per regimen or the demand from outbreaks would have to increase by orders of magnitude to have the same effect. We encourage readers to engage with these assumption parameters critically using our open sourcesoftware.

While the main focus of this paper is the challenge of financing EID vaccine development, we realize that there are other concerns that must be considered in parallel before a portfolio of novel EID vaccine regimens is made available to the public. These issues include, but are not limited to, preclinical discovery, regulatory approval strategy, and post-approval procurement and distribution. These are matters of great importance and warrant further investigation.

It is indisputable, however, that better business models for global health security are urgently needed. We expect there may be benefits to extending the scope of the megafund approach beyond the particular EID vaccine assets considered in this study, perhaps to antibiotics or MCMs for intentional biological threats, an additional global health security concern. While this would do little to improve the desirability of EID vaccine candidates as assets, broadening the scope of a fund to address additional threats may create greater financial viability to global health security more broadly.

As past efforts demonstrate, the key to generating interest in developing vaccine assets is to offer sufficient financial incentives for would-be developers, such as direct market commitments or priority review vouchers. Closing the gap between the economic value of epidemic prevention and the financial returns of vaccine assets, whether by encouraging the market to compensate developers through a capitated vaccine “subscription” model, or by combining vaccine assets into a large portfolio to normalize investment risk as described above, will better enable the global health security community to address the dangers of EIDs.

## Data Availability

All data and software used in this manuscript is publicly available.

## Acknowledgments

We thank Ellen Carlin, Doug Criscitello, Margaret Crotty, Narges Dorratoltaj, Per Etholm, Jeremy Farrar, Nimah Farzan, Mark Feinberg, Jose-Maria Fernandez, John Grabenstein, Peter Hale, Richard Hatchett, Peter Hotez, Daniel Kaniewski, Adel Mahmoud, Mike Osterholm, Kevin Outterson, Chi Heem Wong, CEPI leadership, and two reviewers and the editor for helpful comments and discussion, and Jayna Cummings for editorial assistance. Research support from the MIT Laboratory for Financial Engineering and the Warren Alpert Medical School of Brown University is gratefully acknowledged. The views and opinions expressed in this article are those of the authors only, and do not necessarily represent the views and opinions of any institution or agency, any of their affiliates or employees, or any of the individuals acknowledged above.

## Prior Work

Several mechanisms have recently been proposed or implemented to create incentives for industry to develop vaccines and other medical countermeasures for EIDs *(1)*. These strategies include the direct government acquisition of stockpiles of vaccines, the use of prizes, priority review vouchers, and the establishment of advance market commitments, we describe in more detail below.

### Government Research, Development, and Acquisition

It is clear that direct, non-dilutive funding for R&D will continue to be integral to future vaccine development efforts. Governments, nonprofit organizations such as the Gates Foundation and the Wellcome Trust, and the recently established CEPI are committed to provide this funding. These entities offset the exceptional risk faced by vaccine developers beyond the traditional scientific risk. The operational, regulatory, and market risks of vaccine development remain extraordinary. Without robust and sustained R&D funding, many early-stage assets cannot succeed. While R&D funding “push” mechanisms are necessary, however, they alone are typically not sufficient.

In response to the anthrax attacks of 2001 and the outbreaks of SARS and H5N1 avian influenza in 2003 and 2004, the U.S. government established two programs to address biodefense and pandemic threats. These programs, Project BioShield *(2)* and Pandemic Influenza Preparedness *(3)*, mandated that the U.S. government acquire stockpiles of medical countermeasures (MCMs) against these threats. Each of these preparedness programs was funded with an approximately $6 billion, multi-year appropriation. Central to both programs was the establishment of guaranteed markets to purchase vaccines and other MCMs. This was necessary to mitigate the pandemic threat for the U.S. population, but as importantly, to establish viable public-private partnerships, as the U.S. government does not have licensed vaccine-manufacturing capabilities itself. Stockpiles have since been established for a range of MCMs, and are in progress for others, including vaccines for the Ebola virus *(2, 4–6)*.

### Prizes

Historically, prizes have often been used as an incentive for technological innovation. For example, the first Kremer Prize of £50,000 was awarded in 1977 for the invention of the “first substantial flight of a human-powered airplane” *(7)*. While some experts believe that this approach might create sufficient incentives for research and development in less commercially attractive diseases *(8, 9)*, there is substantial difficulty in applying this structure to EID vaccines, as the prize pool would have to be large enough to offset the high development costs. As a result, several experts have proposed market-based approaches instead *(10, 11)*. Most recently, a prize model has been proposed to incentivize the development of novel antibiotics to address the increasing global problem of antibiotic resistance *(12)*. Importantly, the price is delinked from the volume of sales (as with U.S. government acquisition programs), a key issue for EIDs, where volumes are often insufficient to drive viable markets *(13)*.

### Priority Review Vouchers

Another mechanism is the FDA priority review voucher program, currently implemented by the U.S. government. Under this program, first proposed by Ridley *et al. (11)*, companies developing a therapy for a traditionally “neglected” disease can apply for an FDA priority review voucher. Such vouchers can be used by the company for the accelerated review of another, potentially more lucrative asset, or sold to another firm for review of one of their own assets. Extending this program to medical countermeasures has been under consideration for years *(1)*, and the U.S. 21st Century Cures Act expanded the scope of the program to MCMs for material threats (e.g., smallpox), now including Ebola and Zika *(14, 15)*. An analysis by Berman and Radhakrishna suggests that these vouchers have tremendous value, with one selling for as much as $350 million on the open market *(16)*. However, their value may be waning as more become available, as acquisition prices have decreased over the last few years *(15)*. Even so, the idea has garnered significant attention, and a European equivalent overseen by the European Medicines Agency (EMA) has been proposed *(17)*.

While some see priority review vouchers as a step in the right direction, vouchers are not without potential drawbacks. For example, vouchers do little to ensure that subsequent vaccine development will be pursued once the first candidate has been approved *(8, 15)*. It is also unclear that the resultant vaccines will ultimately reach patients after approval, after the vouchers have been assigned, once market realities are taken into account *(8)*. They provide one-time revenues to a firm, and do little to ensure sustained manufacturing capability or availability of a vaccine. It should also be noted that the FDA priority review may result in a rejection, making the value of the voucher to a firm more variable than it first appears *(15)*.

### Advance Market Commitments

The final mechanism under consideration is the advance market commitment. This concept is similar to the advance purchasing commitments that the U.S. government can make under Project BioShield for MCMs up to eight years in advance of their licensure *(18)*. Advance market commitments allow vaccine developers to assess the potential demand for their product if approved, and provide some guarantee of expected compensation for their efforts. Levine *et al. (10)* describe how such a structure would operate. Essentially, stakeholders from wealthy countries would agree to pay a certain price per dose for a successful vaccine against a target disease, subsidizing the amount that a poorer country would pay, should the development project prove successful. While the risk of scientific failure would still be present, some of the potential demand and revenue would be quantified before the project would be undertaken, serving as encouragement to prospective developers. However, this approach assumes that wealthier entities will still be interested in purchasing vaccines for relatively rare diseases that might not have a direct impact on their constituents *(19)* unless a significant outbreak emerges.

While the methods described here may help mitigate the shortcomings of vaccine investment, they also suggest that a more sustainable long-term solution lies in aligning the incentives for wealthier stakeholders with the incentives of those people most vulnerable to EIDs. Indeed, this alignment is prudent for the former group, as under outbreak conditions their health security may be at risk, even in places where EIDs are unlikely to emerge *(19)*, as the recent Zika and Ebola outbreaks illustrate.

## Flu Pandemics

Recent work by Fan *et al. (20)* calculated that the global expected loss due to pandemic influenza would be approximately $570 billion annually. In 2015, the WHO noted the emergence of many novel influenza viruses, resulting in an “especially volatile” gene pool, left the consequences to human health “unpredictable yet potentially ominous” *(21)*. World Bank projections give a sense of the cost of inaction: a worldwide influenza epidemic would reduce global wealth by an estimated $3 trillion *(22)*. Even with diligent containment efforts and antiviral therapy, Colizza *et al*. suggest that a particularly infectious strain might still infect 30-50% of the global population *(23)*, making prophylactic vaccines essential in mitigating pandemic risk *(24)*.

## Portfolio Simulation Analysis

We present the details of our simulation analysis of the expected risks and returns of a portfolio of 141 preclinical emerging infectious disease (EID) vaccine candidate projects, as well as the assumptions used to estimate the annual expected revenues from direct sales of vaccines to susceptible populations for the 9 different EIDs addressed in our megafund. In our portfolio, we utilize CEPI’s *(25)* targeted EIDs and pipeline research *(26)*, which is based upon the World Health Organization’s R&D Blueprint for epidemic prevention *(27)* (see Table S1). Diseases for which efficacious vaccines have already been approved outright, such as Dengue fever, or provisionally in emergency situations, such as Ebola, were excluded.

**Table S1.** Portfolio of assets 9 target EIDs and the number of projects for each included in the hypothetical portfolio.

| Disease | *n* |
| --- | --- |
| Chikungunya | 20 |
| MERS | 14 |
| SARS | 6 |
| Marburg | 19 |
| Rift Valley Fever | 15 |
| Lassa Fever | 22 |
| Nipah Virus | 11 |
| Crimean Congo Hemorrhagic Fever | 6 |
| Zika Virus | 28 |

We begin with a discussion of the correlation assumptions underlying the Monte Carlo simulation of our EID megafund portfolio’s performance, and provide details regarding our projected development costs and phase-transition assumptions. We then turn to how investment returns are defined, and conclude by describing our projected revenue estimates for EID vaccines.

### 1. Simulating Correlated Vaccine Candidate Projects

While there are a number of methods for modeling the outcome of clinical trials with certain scientific elements in common, we numerically estimate the performance of our EID vaccine portfolio by modeling projects as pairwise correlated Bernoulli trials. Our methods are similar to Lo *et al. (28)*.

Denote by ϵ ≡ [*ϵ*_1_ *ϵ*_2_ … *ϵ*_*n*_]*′* a column-vector of random multivariate standard normal variables. Then for any positive-definite matrix Σ, the new vector of random variables *Z* = Σ^1/2^ϵ is multivariate normal with covariance matrix Σ, where Σ^1/2^ denotes the Cholesky factorization or matrix square root of Σ. Once the success probability, *p*_*i*_, for each Bernoulli trial random variable *B*_*i*_ is defined, *B*_*i*_ can be simulated as

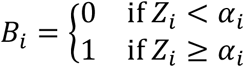

where we define *α*_*i*_ = Φ^-1^(1 - *p*_*i*_) and Φ^-1^(·) is the inverse of the standard normal cumulative distribution function.

For our purposes, pairwise correlations are meant to capture commonalities among translational vaccine development programs, so that success or failure in one program has predictive power for the success or failure of another program. In addition to specifying values for each entry in Σ that are based on domain-specific knowledge of the underlying science, we must also ensure that Σ is a valid positive-definite covariance matrix.

In our simulations, we adopt a three-step process in which all pairwise correlations between projects are first evaluated qualitatively as “low” or “high.” These assessments are then translated into numerical values of 10% for “low” and 50% for “high.” The outline of the dimensions used to assign correlation levels is displayed in Table S2 below. Figure S1 shows a heat map of these assumed correlations.

**Figure S1.**
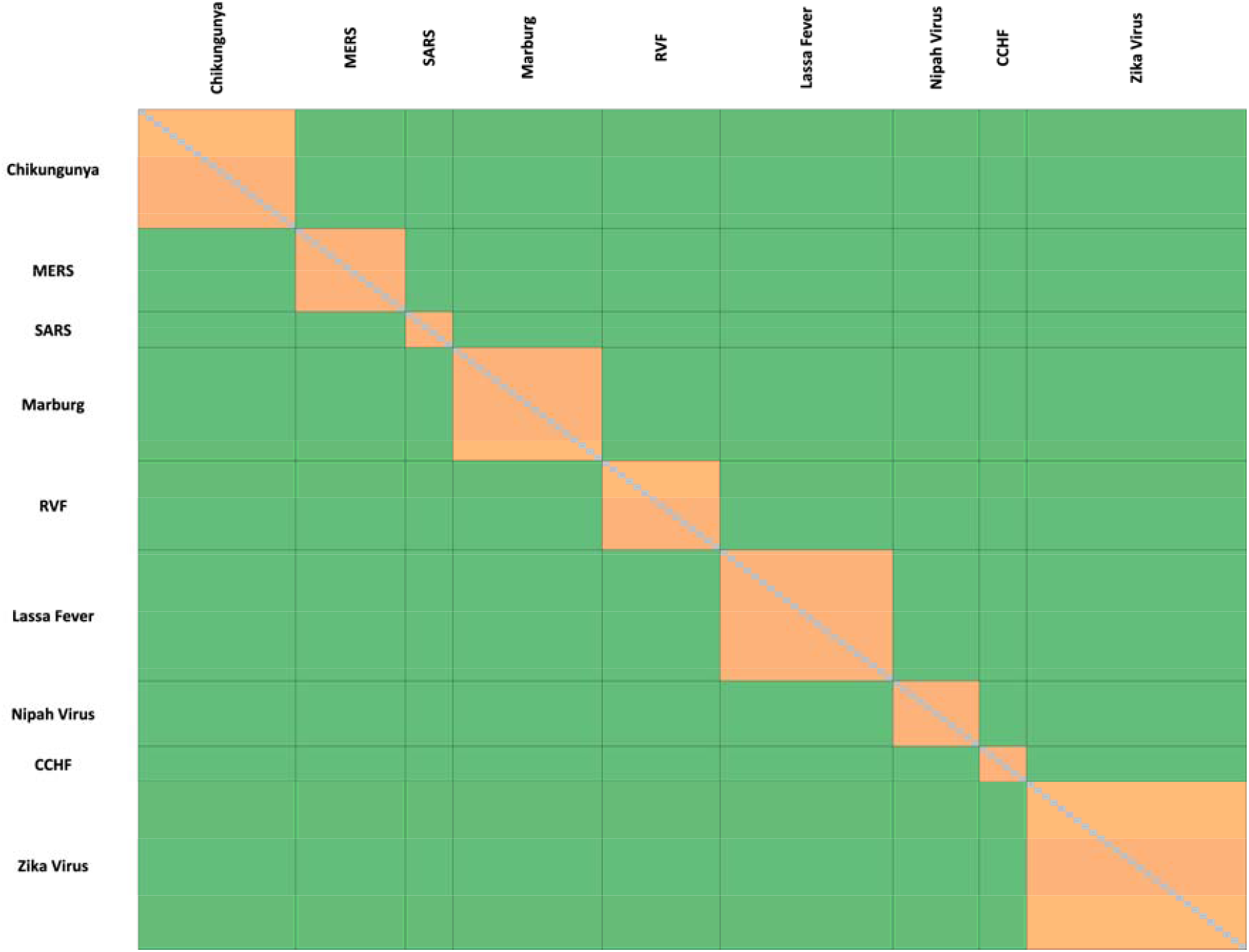
Correlation between vaccine projects. Heat-map representation of qualitatively determined pairwise correlation of success among 141 EID vaccine development projects. Orange cells indicate 50% and green cells indicate 10%

The third step is to apply the numerical algorithm developed by Qi and Sun *(29)* to compute the closest positive-definite matrix to the one specified manually. In this case, the manually defined correlation matrix shown in Figure 2 in the main text was already positive-definite, indicating that the Qi and Sun algorithm had no impact.

**Table S2.** Pairwise correlation assignments based on Targeted Disease

| Disease | Correlation |
| --- | --- |
| Same | High (50%) |
| Different | Low (10%) |

### 2. Development Times, Transition Probabilities, and Research Costs

We use the CEPI estimates of phase-transition probability and development time at each phase in our simulation (shown in Table S3), seeking to develop each asset through phase 2 *(25)*. CEPI assumes that the measures taken by global actors in response to the recent Ebola outbreaks indicate that phase 2 development would justify the stockpiling and conditional use of these candidate vaccines in an actual outbreak setting, one that could support later vaccine approval and distribution *(25)*. We use CEPI’s estimate that the cost to develop each preclinical asset through phase 2 is $250 million.

Our simulation assumes trials with a standard progression from phase to phase. If earlier stages of R&D are included, or if a trial must be repeated, the costs and duration will increase, and the post-approval patent life of the asset will decrease. On the other hand, because we have not modeled the transition from one clinical phase to the next, the realized out-of-pocket cost of a typical project could be less than the assumed $250 million because of the early termination of failed projects. CEPI’s assumption of $250 million of out-of-pocket costs falls well within industry estimates of the vaccine development costs through phase 2 with limited manufacturing scale. Though not analyzed here, the inclusion of phase 3 and manufacturing facility maintenance and surge/scale-up can be factored into the model using our open-source software.

**Table S3.** CEPI’s *(25)* estimated phase-transition probabilities and development time at each phase.

| Phase | Development Time (Years) | Transition Probability (%) |
| --- | --- | --- |
| Preclinical | 1.5 | 57 |
| Phase 1 | 2.0 | 72 |
| Phase 2 | 1.5 | 79 |

### 3. Computing Returns

An investment rate of return, *R*, where an initial investment of *I* yields a single payoff *X* is defined as *R* = (*X*/*I*) - 1. If the investment is over a duration *T* > 1 year, the return is often annualized to simplify comparisons with other investments of different durations. This geometric compounding assumes that interim gains are reinvested, and hence additional interest is paid on the interest earned. The annualized return, *R*_*a*_, is defined as

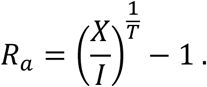

This definition is relatively straightforward. However, a question arises in the computation of expected returns and standard deviations for multi-year returns, which require annualization: should the moments be computed before or after annualization? In the main text, we annualize realized returns before calculating statistics such as expectation and standard deviation. While there is no clear argument for using one method over the other in all contexts, we have chosen to annualize first to calculate the realized internal rate of return (IRR), and then to compute the expected IRR and standard deviation of IRR, which are the more traditional summary statistics.

### 4. Projected Revenues

It is well recognized that predicting the type, frequency, and scale of any future EID outbreak, epidemic, or pandemic with accuracy is not possible, and therefore certain practical assumptions were necessary to project revenues. In our model, the probability of a given disease having an outbreak in a given year is given by the ratio of the number of historical outbreaks to the number of years since the disease was first reported or since the first notable outbreak. Respective probabilities are listed in Table S4 below. This represents a pragmatic approach, and is not expected to reflect actual future epidemiological patterns. While more sophisticated models are available and have been used to support other pandemic financing programs *(30)*, this approach is intended to provide a baseline assessment of megafund financing *(31)*.

**Table S4.**
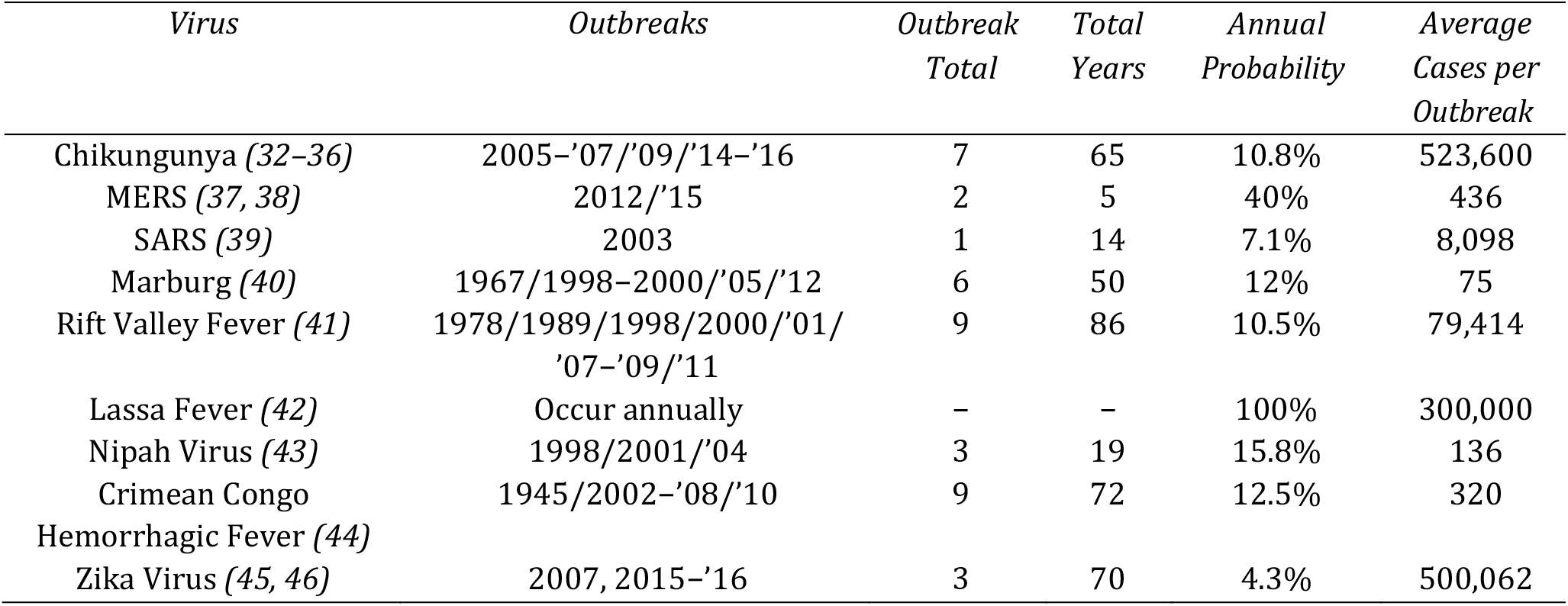
Historical Outbreak Data for Portfolio Diseases *(32, 33, 42–46, 34–41)*.

The number of vaccine regimens sold in response to an outbreak is a function of both the actual and perceived risk to populations. This is subject to significant uncertainty, due in part to gaps in knowledge at the onset of an outbreak about its transmission patterns and its medical and public health impact *(19, 47–49)*. We based our projections of the number of vaccine regimens sold on the average number of infections observed per outbreak, and further modulated by three factors: Woolhouse Potential for Pandemic Spread, Severity of Clinical Symptoms, and Mortality Rate. The number of vaccine regimens sold is given by the average number of cases per outbreak multiplied by Woolhouse weighting and clinical severity rating as described below. This is used as a crude proxy for demand extending beyond those immediately affected, e.g., the so-called ‘worried well’.

By the nature of the methodology that CEPI used to establish their priority list of vaccines, all of the viruses addressed in our portfolio are known to be potent, contagious pathogens. However, the transmissibility between humans will vary. Woolhouse *et al. (50, 51)* categorize EIDs into four levels, described in Table S5 below. Consistent with the criteria used by WHO and CEPI to identify priority pathogens, all of the diseases in our EID portfolio are either Level 3 or Level 4. We assigned Level 3 diseases a weight of 1.0, and Level 4 diseases a weight of 2.0, essentially doubling the vaccine regimens sold relative to those for Level 3 viruses.

**Table S5.** Levels of transmissibility as categorized by Woolhouse *et al*. *(50, 51)*, which are used in the calculation of the number of vaccines sold. All EIDs in the portfolio are level 3 or 4; level 3 diseases in the portfolio are assigned a weight of 1.0 and level 4 diseases in the portfolio are assigned a weight of 2.0.

| Woolhouse Level | Interpretation |
| --- | --- |
| Level 1 | Humans exposed but not infected |
| Level 2 | Humans infected |
| Level 3 | Human-to-human transmission |
| Level 4 | Increased potential for epidemics/persist as endemic infection |

Risks to those not involved in the initial outbreak, both real and perceived, will also drive the demand for these vaccine regimens. As can be seen by the ongoing discussions between the U.S. government and Sanofi regarding licenses of Zika vaccines, any projections of future demand are theoretical at best *(52)*. However, it is difficult to discount the effect of public perception on the willingness of people and policymakers to take action against these pathogens, as illustrated by the discovery of the connection between Zika and congenital microcephaly.

Though all the EIDs studied are a significant threat to human health, each disease presents itself with different symptoms and a unique prognosis. These differences in presentation and outcome may affect the way in which the public responds to outbreaks. We rate each disease by clinical presentation and mortality rate as mild, mild-moderate, moderate, moderate-severe or severe in Table S6, and assign a corresponding multiplier in Table S7. The multipliers in Table S7 were informed by recent developments about a promising new vaccine candidate for Ebola. According to the most recent reports, Merck has promised to produce 300,000 doses of the vaccine *(53)*, while the 2015 outbreak totaled approximately 30,000 cases *(54)*. Given the severity of clinical symptoms and high mortality rate associated with Ebola infection, we assign a multiplier of 10 to our “severe” category of diseases, and adjust our multiplier accordingly based on clinical severity. While each of these diseases has the potential to cause severe illness, some are asymptomatic in most patients, and thus less likely to elicit high demand for a resulting vaccine. In assigning ratings, we also assumed that the potential for certain sequelae will increase demand for certain vaccines; while the presentation of Zika is generally mild, the possibility of birth defects resulting from infection in pregnant women will likely boost the demand for this vaccine, increasing its relative rating.

**Table S6.** Rating of each disease by clinical presentation and mortality.

| Disease | Clinical Severity Rating |
| --- | --- |
| Chikungunya | Mild |
| MERS | Severe |
| SARS | Moderate |
| Marburg | Severe |
| Rift Valley Fever | Moderate |
| Lassa Fever | Mild-Moderate |
| Nipah Virus | Severe |
| Crimean Congo Hemorrhagic Fever | Severe |
| Zika Virus | Moderate |

**Table S7.** Corresponding demand multipliers for each clinical severity rating.

| Clinical Severity Rating | Multiplier |
| --- | --- |
| Mild | 2x |
| Mild-Moderate | 4x |
| Moderate | 6x |
| Moderate-Severe | 8x |
| Severe | 10x |

The price per dose of vaccine was estimated by taking the average of prices listed in the CDC Adult vaccine price list 2016 *(55)*; UNICEF’s 2016 product menu for Gavi, the Vaccine Alliance *(56)*; and the Pan American Health Organization (PAHO) expanded program of immunization vaccine prices for 2016 *(57)*. These three averages serve as our vaccine price for high, low, and middle income countries respectively. We then use these to price each vaccine based on the income level of the countries most likely to have an outbreak of a particular disease. (The endemic country/disease/pricing per dose pairings are listed in Table S8 below.) It should be noted that we took a conservative stance on pricing in our model, opting to model the lower income country price for diseases that have historically emerged in nations with differing ability to pay. Our pricing is also likely conservative due to our inclusion of each vaccine on each menu in our mean calculations, including older vaccines that are apt to be produced and sold at lower cost than new vaccines.

The annual expected revenue for each vaccine candidate is then given by the price per dose of vaccine times the expected number of vaccines sold in an outbreak weighted by the probability of an outbreak occurring in a given year.

**Table S8.**
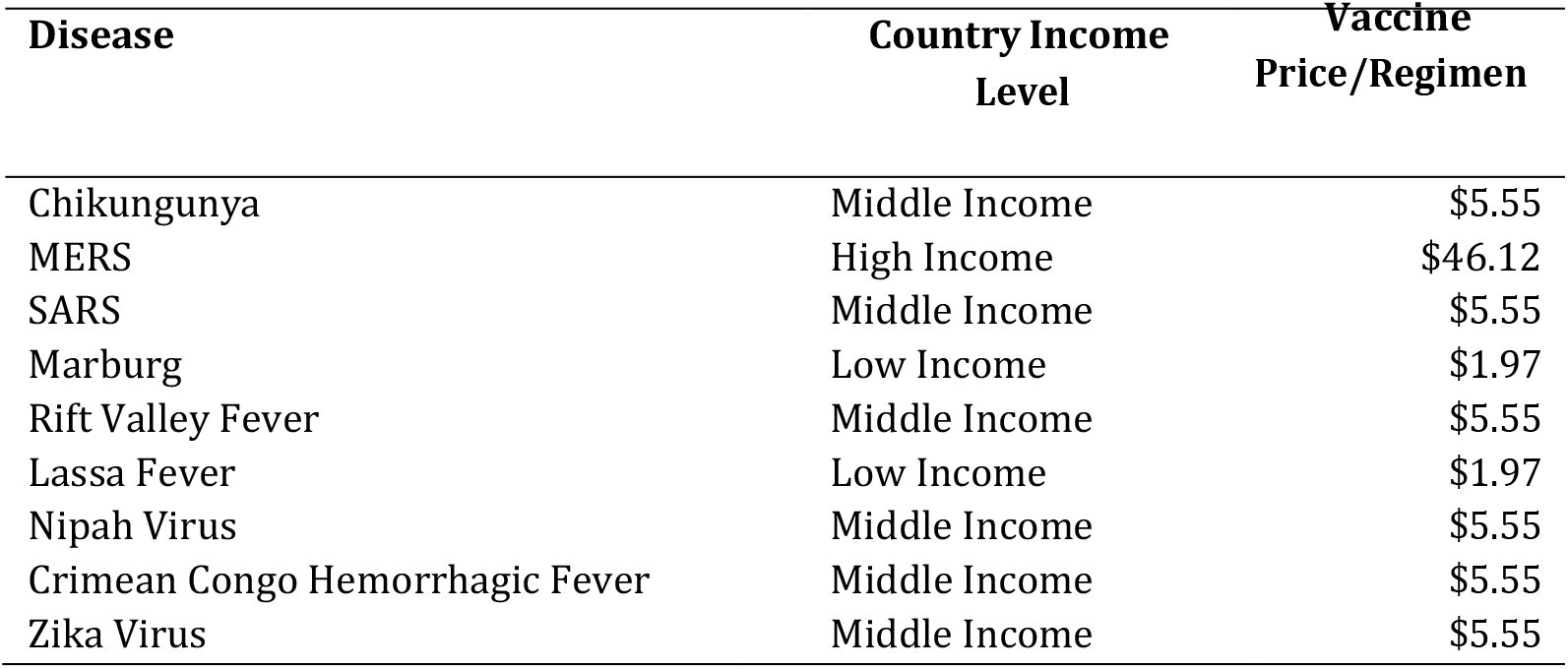
Vaccine price per dose assumptions based on the income level of the countries most likely to have an outbreak of each disease included in the EID portfolio.

### 5. Sensitivity Analysis

This simulation parameterizes several assumptions about the cost and duration of vaccine development, the probability of success, and pairwise correlations of success between the projects. These estimates were based on the published literature on vaccine development and qualitative input from scientists with domain-specific expertise.

In this section, we investigate the robustness of our results to the parameterized assumptions of our model. We update the investment return statistics of the EID vaccine portfolio as we vary the development cost and probability of success of each project. The expected return and return standard deviation associated with the perturbed parameters are given in Tables S9 and S10.

In Table S9, we find that the expected return of the portfolio increases as the cost per project decreases. Similarly, Table S10 reports that the expected return of the portfolio increases as the probability of success of each project increases. However, even under more optimistic assumptions, the expected annualized return of the megafund for the base case remains significantly negative, increasing from –61.1% to only –57.4% when the probability of success is increased by 150%. This sensitivity analysis underscores the robustness of our results, and demonstrates that an EID vaccine portfolio remains economically unviable even under relatively optimistic cost and revenue assumptions.

Finally, Table S11 considers the performance of the EID vaccine portfolio under the scenario where a government agency or philanthropic organization agrees to absorb the initial losses on the portfolio for a predetermined amount, which we specify as 25% and 50% of our simulated megafund’s principal. We find that, under the base case scenario, the expected return increases from –61.1% to –23.6% and –12.6%, respectively. While these scenarios remain unprofitable, it demonstrates that if combined with other revenue-boosting mechanisms such as such as advance market commitments and priority review vouchers, the guarantee structure has the potential to transform an unattractive portfolio of EID vaccine candidates into one that could realistically attract private-sector capital.

**Table S9.**
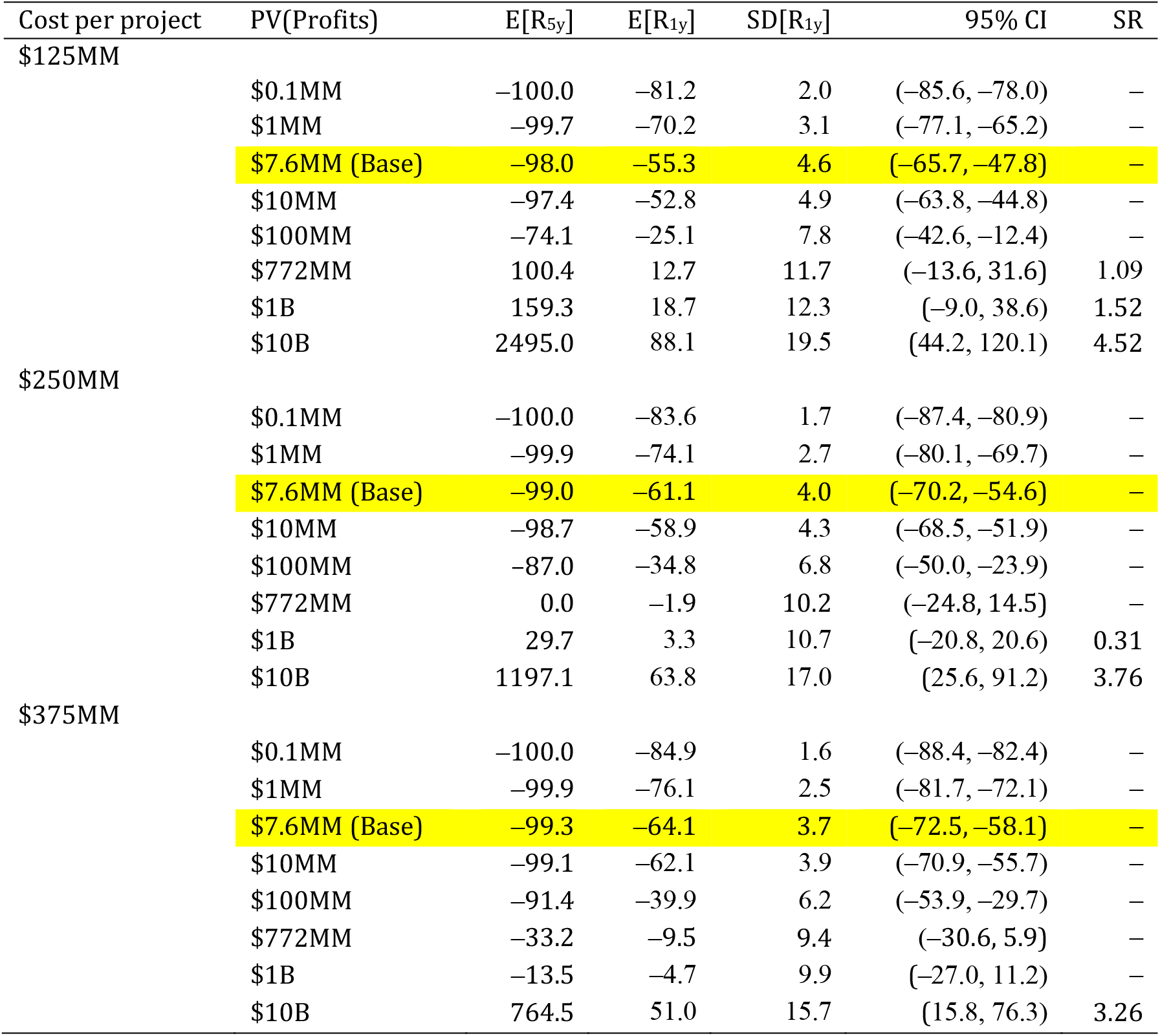
Sensitivity of the investment returns (%) of a portfolio of 141 preclinical EID vaccine candidates to project development costs. The table reports the results for [50%, 100%, 150%] of the $250MM cost per project proposed in the study. The Sharpe ratio is estimated as the ratio of the expected return to the standard deviation. PV(Profits), present value of profits per successful vaccine in year 5; E[R_5y_], expected 5-year return on investment; E[R_1y_], expected annualized return; SD[R_1y_], annualized return standard deviation; CI, confidence interval; SR, Sharpe ratio.

| Cost per project | PV(Profits) | E[R<sub>5y</sub>] | E[R<sub>1y</sub>] | SD[R<sub>1y</sub>] | 95% CI | SR |
| --- | --- | --- | --- | --- | --- | --- |
| \$125MM | | | | | | |
| | \$0.1MM | -100.0 | -81.2 | 2.0 | (-85.6, -78.0) | - |
| | \$1MM | -99.7 | -70.2 | 3.1 | (-77.1, -65.2) | - |
| | **\$7.6MM (Base)** | **-98.0** | **-55.3** | **4.6** | **(-65.7, -47.8)** | **-** |
| | \$10MM | -97.4 | -52.8 | 4.9 | (-63.8, -44.8) | - |
| | \$100MM | -74.1 | -25.1 | 7.8 | (-42.6, -12.4) | - |
| | \$772MM | 100.4 | 12.7 | 11.7 | (-13.6, 31.6) | 1.09 |
| | \$1B | 159.3 | 18.7 | 12.3 | (-9.0, 38.6) | 1.52 |
| | \$10B | 2495.0 | 88.1 | 19.5 | (44.2, 120.1) | 4.52 |
| \$250MM | | | | | | |
| | \$0.1MM | -100.0 | -83.6 | 1.7 | (-87.4, -80.9) | - |
| | \$1MM | -99.9 | -74.1 | 2.7 | (-80.1, -69.7) | - |
| | **\$7.6MM (Base)** | **-99.0** | **-61.1** | **4.0** | **(-70.2, -54.6)** | **-** |
| | \$10MM | -98.7 | -58.9 | 4.3 | (-68.5, -51.9) | - |
| | \$100MM | -87.0 | -34.8 | 6.8 | (-50.0, -23.9) | - |
| | \$772MM | 0.0 | -1.9 | 10.2 | (-24.8, 14.5) | - |
| | \$1B | 29.7 | 3.3 | 10.7 | (-20.8, 20.6) | 0.31 |
| | \$10B | 1197.1 | 63.8 | 17.0 | (25.6, 91.2) | 3.76 |
| \$375MM | | | | | | |
| | \$0.1MM | -100.0 | -84.9 | 1.6 | (-88.4, -82.4) | - |
| | \$1MM | -99.9 | -76.1 | 2.5 | (-81.7, -72.1) | - |
| | **\$7.6MM (Base)** | **-99.3** | **-64.1** | **3.7** | **(-72.5, -58.1)** | **-** |
| | \$10MM | -99.1 | -62.1 | 3.9 | (-70.9, -55.7) | - |
| | \$100MM | -91.4 | -39.9 | 6.2 | (-53.9, -29.7) | - |
| | \$772MM | -33.2 | -9.5 | 9.4 | (-30.6, 5.9) | - |
| | \$1B | -13.5 | -4.7 | 9.9 | (-27.0, 11.2) | - |
| | \$10B | 764.5 | 51.0 | 15.7 | (15.8, 76.3) | 3.26 |

**Table S10.**
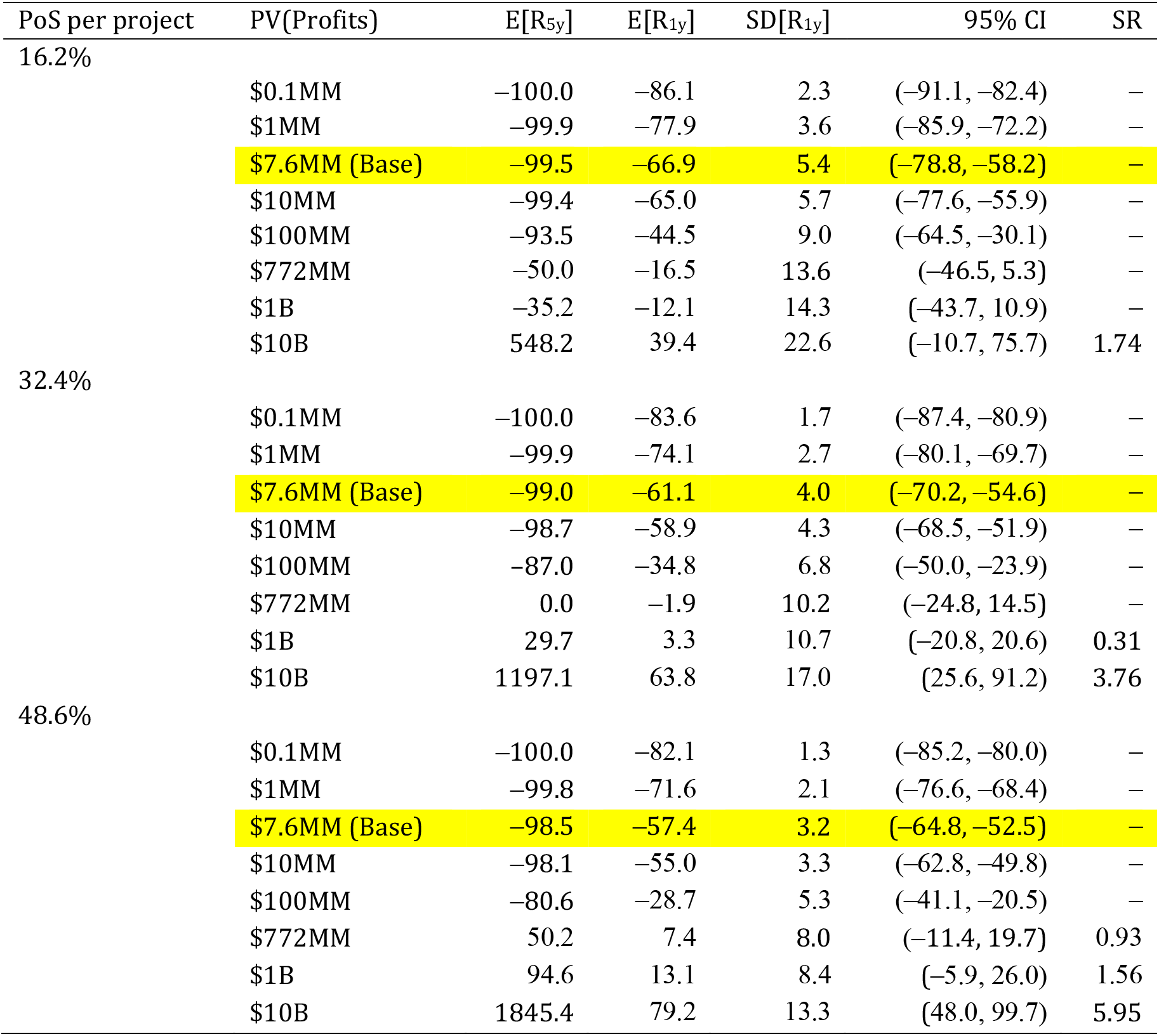
Sensitivity of the investment returns (%) of a portfolio of 141 preclinical EID vaccine candidates to development success rates. The table reports the results for [50%, 100%, 150%] of the 32.4% probability of success (PoS) estimate for each project proposed in the study. The Sharpe ratio is estimated as the ratio of the expected return to the standard deviation. PV(Profits), present value of profits per successful vaccine in year 5; E[R_5y_], expected 5-year return on investment; E[R_1y_], expected annualized return; SD[R_1y_], annualized return standard deviation; CI, confidence interval; SR, Sharpe ratio.

**Table S11.** Sensitivity of the investment returns (%) of a portfolio of 141 preclinical EID vaccine candidates under a government-backed guarantee structure. The table reports the results for a guarantee on [0%, 25%, 50%] of the portfolio’s principal proposed in the study. The Sharpe ratio is estimated as the ratio of the expected return to the standard deviation. PV(Profits), present value of profits per successful vaccine in year 5; E[R_5y_], expected 5-year return on investment; E[R_1y_], expected annualized return; SD[R_1y_], annualized return standard deviation; CI, confidence interval; SR, Sharpe ratio.

| %Guarantee | PV(Profits) | E[R<sub>5y</sub>] | E[R<sub>1y</sub>] | SD[R<sub>1y</sub>] | 95% CI | SR |
| --- | --- | --- | --- | --- | --- | --- |
| 0% | \$0.1MM | -100.0 | -83.6 | 1.7 | (-87.4, -80.9) | - |
| | \$1MM | -99.9 | -74.1 | 2.7 | (-80.1, -69.7) | - |
| | **\$7.6MM (Base)** | **-99.0** | **-61.1** | **4.0** | **(-70.2, -54.6)** | **-** |
| | \$10MM | -98.7 | -58.9 | 4.3 | (-68.5, -51.9) | - |
| | \$100MM | -87.0 | -34.8 | 6.8 | (-50.0, -23.9) | - |
| | \$772MM | 0.0 | -1.9 | 10.2 | (-24.8, 14.5) | - |
| | \$1B | 29.7 | 3.3 | 10.7 | (-20.8, 20.6) | 0.31 |
| | \$10B | 1197.1 | 63.8 | 17.0 | (25.6, 91.2) | 3.76 |
| 25% | \$0.1MM | -75.0 | -24.2 | 0.0 | (-24.2, -24.2) | - |
| | \$1MM | -74.9 | -24.1 | 0.0 | (-24.2, -24.1) | - |
| | **\$7.6MM (Base)** | **-74.0** | **-23.6** | **0.3** | **(-24.1, -23.1)** | **-** |
| | \$10MM | -73.7 | -23.5 | 0.3 | (-24.0, -22.7) | - |
| | \$100MM | -62.0 | -17.8 | 2.5 | (-22.4, -12.7) | - |
| | \$772MM | 10.9 | 1.2 | 6.7 | (-13.3, 14.8) | 0.18 |
| | \$1B | 36.2 | 5.2 | 8.0 | (-10.9, 20.6) | 0.65 |
| | \$10B | 1197.4 | 63.8 | 16.9 | (25.6, 91.2) | 3.78 |
| 50% | \$0.1MM | -50.0 | -12.9 | 0.0 | (-12.9, -12.9) | - |
| | \$1MM | -49.9 | -12.9 | 0.0 | (-12.9, -12.9) | - |
| | **\$7.6MM (Base)** | **-49.0** | **-12.6** | **0.2** | **(-12.9, -12.3)** | **-** |
| | \$10MM | -48.7 | -12.5 | 0.2 | (-12.8, -12.1) | - |
| | \$100MM | -37.0 | -8.9 | 1.7 | (-11.9, -5.5) | - |
| | \$772MM | 16.5 | 2.6 | 5.0 | (-5.8, 14.5) | 0.52 |
| | \$1B | 39.5 | 6.0 | 6.9 | (-4.1, 20.6) | 0.87 |
| | \$10B | 1196.7 | 63.8 | 16.9 | (25.6, 91.2) | 3.78 |

## Acknowledgments

We thank Ellen Carlin, Doug Criscitello, Narges Dorratoltaj, Per Etholm, Jeremy Farrar, Nimah Farzan, Mark Feinberg, Jose-Maria Fernandez, John Grabenstein, Peter Hale, Richard Hatchett, Peter Hotez, Daniel Kaniewski, Adel Mahmoud, Kevin Outterson, Chi Heem Wong, and CEPI leadership for helpful comments and discussion, and Jayna Cummings for editorial assistance. Research support from the MIT Laboratory for Financial Engineering is gratefully acknowledged. The views and opinions expressed in this article are those of the authors only, and do not necessarily represent the views and opinions of any institution or agency, any of their affiliates or employees, or any of the individuals acknowledged above.

